# Clinical Outcomes in Hospitalized Atrial Tachyarrhythmia Patients With and Without Prior Thoracic Irradiation

**DOI:** 10.1101/2025.09.20.25336164

**Authors:** Aren Singh Saini, Sahil Ghay, Baneet Kaur, Shalina Chithriki, Pooja Ghay, Raksha Narasimhan, Kayla Samimi, Isabella Dreyfuss, Roopin Pal Singh, Brandon Mahal, Crystal Seldon Taswell

**Affiliations:** Department of Radiation Oncology, Sylvester Comprehensive Cancer Center, University of Miami Miller School of Medicine, Miami, FL, USA; Department of Internal Medicine, Mount Sinai Medical Center, Miami Beach, FL, USA; Edward Via College of Osteopathic Medicine, Monroe, LA, USA

**Keywords:** Atrial Arrhythmias, Atrial Fibrillation, Atrial Flutter, Radiation Therapy, Cancer, Cardio-Oncology, Outcomes

## Abstract

**Background:** Thoracic radiation therapy (TRT) is commonly used for breast, lung, and lymphoid cancers. While its cardiotoxic effects, particularly coronary artery disease, are well recognized, less is known about its impact on arrhythmia-related hospitalizations.

**Methods:** A retrospective cohort study using the National Inpatient Sample (2016–2022) was conducted. Admissions for atrial fibrillation or flutter were identified using ICD-10 codes, and prior TRT was determined from thoracic malignancy and radiation history codes. Propensity score matching and doubly robust multivariable regression were used to evaluate outcomes. The primary endpoint was in-hospital mortality; secondary endpoints included length of stay (LOS) and total charges.

**Results:** Among 3,198,304 weighted admissions, 8,570 (0.27%) had prior TRT. After matching, TRT was associated with higher odds of in-hospital mortality (adjusted odds ratio [aOR] 1.97; 95% CI 1.17– 3.32; p=0.010) and longer LOS (+0.30 days; 95% CI 0.05–0.55; p=0.019) without increased costs (p=0.202). TRT patients also had higher odds of palliative consultation (aOR 2.60, p<0.001) and DNR status (aOR 1.97, p<0.001), but lower odds of acute kidney injury (aOR 0.66, p<0.001).

**Conclusion:** Prior TRT is linked to greater in-hospital mortality and resource utilization during atrial fibrillation or flutter admissions, likely reflecting cumulative cardiovascular injury. These findings support closer surveillance and early intervention for this high-risk population.

## Introduction

Thoracic irradiation is central to the treatment of breast, lung, and lymphoid cancers. As more patients live longer after treatment, late cardiovascular toxicity has become an important long-term health concern. Radiation-induced heart disease includes coronary, valvular, pericardial, and myocardial injury. Radiation may also remodel atrial tissue and the cardiac conduction system. These changes can create an arrhythmogenic substrate that emerges years after therapy.

Radiation injury arises from endothelial dysfunction, microvascular rarefaction, chronic inflammation, and fibrosis. In the coronary circulation, risk rises with dose. Each 1 Gy increase in mean heart dose is associated with an approximately 7% relative increase in major coronary events, and risk persists for decades [1]. Many late effects emerge 5 to 20 years after exposure [1]. Fibrotic remodeling can produce valvular stenosis or regurgitation, constrictive pericarditis, and restrictive cardiomyopathy. When injury involves the atria or the conduction system, particularly the sinoatrial node and the junction of the left atrium and the pulmonary veins, it predisposes patients to atrial fibrillation (AF) and atrial flutter [2].

Because AF and atrial flutter are the most common sustained arrhythmias in adults, even a modest radiation-related risk could impose substantial inpatient burden. In the United States in 2021, there were 442,909 hospital discharges with a principal diagnosis of AF or atrial flutter [3].

Both bradyarrhythmia and atrial tachyarrhythmia have been documented after thoracic irradiation. Conduction system disease appears more frequent in previously irradiated patients, consistent with fibrosis of the sinus and atrioventricular nodes [2, 4]. In a retrospective lung cancer cohort treated with definitive chemoradiotherapy, a higher maximum sinoatrial node dose was independently associated with increased risk of new-onset atrial fibrillation and with worse overall survival [5]. Similarly, higher left atrial and pulmonary vein doses have been correlated with greater atrial fibrillation incidence in thoracic radiotherapy cohorts [2, 6].

These observations support a dose– and location-dependent risk for atrial arrhythmias, but whether a prior thoracic radiation history alters short-term outcomes once atrial fibrillation or flutter leads to hospitalization remains poorly defined. Existing studies have largely focused on incidence of arrhythmia occurrence rather than prognosis during arrhythmia-related admissions [2, 7]. Most are constrained by limited sample sizes, single-institution settings, and variable eras of radiation delivery, which hinder robust inference regarding in-hospital mortality, length of stay, and resource utilization.

These outcomes capture the immediate clinical burden of arrhythmia admissions and reflect decisions that may be influenced by radiation-related structural heart disease. Mortality risk may be compounded by the interaction of atrial arrhythmias with radiation-associated coronary artery disease, valvular dysfunction, and pulmonary comorbidity [8, 9]. Length of stay and hospital charges may be affected by rhythm control and anticoagulation complexities in oncology populations, as well as differences in procedural utilization and discharge planning among previously irradiated patients. Recognizing that age, comorbidities, and cancer-related factors may confound these relationships underscores the importance of propensity score matching and multivariable adjustment in such analyses.

We therefore examined a contemporary national cohort of atrial fibrillation and atrial flutter admissions to test whether a documented history of thoracic radiation is associated with differences in in-hospital outcomes after matching and doubly robust adjustment for clinical covariates. Using a large, all-payer database and International Classification of Diseases, Tenth Revision (ICD-10) coding, we evaluated whether a history of thoracic irradiation is associated with differences in in-hospital mortality, length of stay, total hospital charges, and various outcomes among adults admitted with atrial fibrillation or flutter. This approach provides broad generalizability and complements substructure dose–response studies by addressing the real-world inpatient prognosis of patients with prior thoracic irradiation.

## Methods

From the Agency for Healthcare Research and Quality (AHRQ), the National Inpatient Sample (NIS) is one of the largest databases available, representing data from non-government-owned short□term, acute□care hospitals in the United States. It is a part of the larger Healthcare Cost and Utilization Project. The NIS is released annually, comprises a stratified, nationally representative sample of approximately 20% of all hospital discharges in the U.S. Discharge weights provided by the NIS can be applied to produce estimates that reflect national trends. Information in NIS is encoded by the International Classification of Diseases, Tenth Revision, Clinical Modification and Procedure Coding System (ICD-10-CM/ICD-10-PCS). Information contained in the dataset includes patient-level demographics, diagnoses, procedures, outcomes, and hospitalization costs. All data in the NIS is de-identified and publicly available; thus, this project is exempt from institutional review board approval.

Data from the NIS (2016–2022) were used to conduct a retrospective cohort study examining inpatient hospitalizations over this period. The STATA18 application was used for all data analysis. All patients 18 and older at the time of admission were identified. Using ICD-10-CM codes beginning with I48, all patients who were admitted with a primary diagnosis of atrial fibrillation or atrial flutter were identified. ICD-10-CM code Z92.3 was used in combination with ICD-10-CM codes for thoracic malignancies to discern which patients received prior thoracic irradiation [Supplement 1]. Baseline characteristics were collected for patients with and without a history of thoracic irradiation including age, gender, race, ethnicity, financial identifiers, comorbidities and hospital characteristics such as hospital location, size and teaching status.

To account for potential confounding, we performed 1:1 nearest-neighbor propensity score matching (PSM) without replacement, applying a caliper of 0.2. Propensity scores were calculated using multivariable logistic regression incorporating demographic, hospital, and clinical variables. These included age, race/ethnicity, gender, ZIP code–based income quartile, insurance type, Charlson comorbidity index category, and hospital characteristics (bed size, region, and teaching status). Clinical covariates included alcohol use, chronic obstructive pulmonary disease, congestive heart failure, diabetes, hepatic disease, hyperlipidemia, hypertension, peripheral vascular disease, pulmonary circulation disorders, including pulmonary hypertension, prior chemotherapy exposure, and valvular heart disease. Covariate balance post-matching was assessed via standardized mean differences and variance ratios, confirming adequate matching quality (mean difference of 2.1% with all standardized mean differences <10%).

A doubly robust regression model was used to analyze the matched cohort to reduce residual confounding with multivariable logistic and linear regression models. Binary outcomes were analyzed using multivariable logistic regression, while continuous outcomes were evaluated with linear regression. Patients with incomplete data were excluded from the analysis. All models were adjusted for the same baseline covariates included in the original propensity score specification. P-values below 0.05 were interpreted as statistically significant.

Two analyses were performed. The primary analysis compared patients with a history of thoracic irradiation to those without among all hospitalizations for atrial tachyarrhythmias. The secondary analysis compared patients treated with radiation therapy only to those treated with chemotherapy only. A separate propensity score model using the same covariates was applied, with 1:1 matching performed without replacement. Binary and continuous outcomes were evaluated. Mortality was the primary outcome, and secondary outcomes included resource utilization metrics (length of stay, total charges) and clinical outcomes such as acute kidney injury, blood transfusion, cardiac arrest, CPR, DNR status, ischemic stroke, mechanical circulatory support, mechanical ventilation, palliative consultation, and vasopressor use.

To evaluate whether the association between prior thoracic radiation and clinical or resource outcomes differed by tumor type, we constructed fully adjusted multivariable regression models for each prespecified endpoint: in-hospital mortality, ischemic stroke, acute kidney injury, cardiac arrest, cardiopulmonary resuscitation, blood transfusion, vasopressor use, mechanical circulatory support, mechanical ventilation, palliative care consultation, do-not-resuscitate status, total hospital costs, and length of stay. Each model included an interaction term between prior thoracic radiation and tumor type. A global Wald test was used to determine whether tumor type significantly modified the effect of prior radiation. For tumor categories with limited sample size that precluded stable model estimation, we repeated analyses using broader tumor groupings to ensure robustness. Statistical significance for interaction was defined as a two-sided p < 0.05.

## Results

From 2016 to 2022, a total of 639,661 patients hospitalized for atrial tachyarrhythmias were identified, of whom 1,714 had a documented history of thoracic radiation therapy. After applying survey weights, this corresponded to an estimated 3,198,304 hospitalizations nationally, including 8,570 (0.27%) with prior thoracic irradiation. Compared to those without prior thoracic irradiation, patients with a history of thoracic radiation were older on average (72.5 vs 70.8 years, p<0.001) but no significant differences in gender were seen (52.9% vs 51.3%, p=0.19) [Table 1]. Significant differences in racial distribution were observed, with a higher proportion of White patients in the irradiated group (82.2% vs 79.9%, p<0.001) and a lower proportion of Hispanic and Asian patients [Table 1]. Primary payment type differed between both groups, with a greater share of patients in the thoracic irradiation group insured by Medicare (76.0% vs 69.0%, p<0.001), and fewer with private insurance (14.2% vs 19.9%, p<0.001) [Table 1].

**Table 1.**
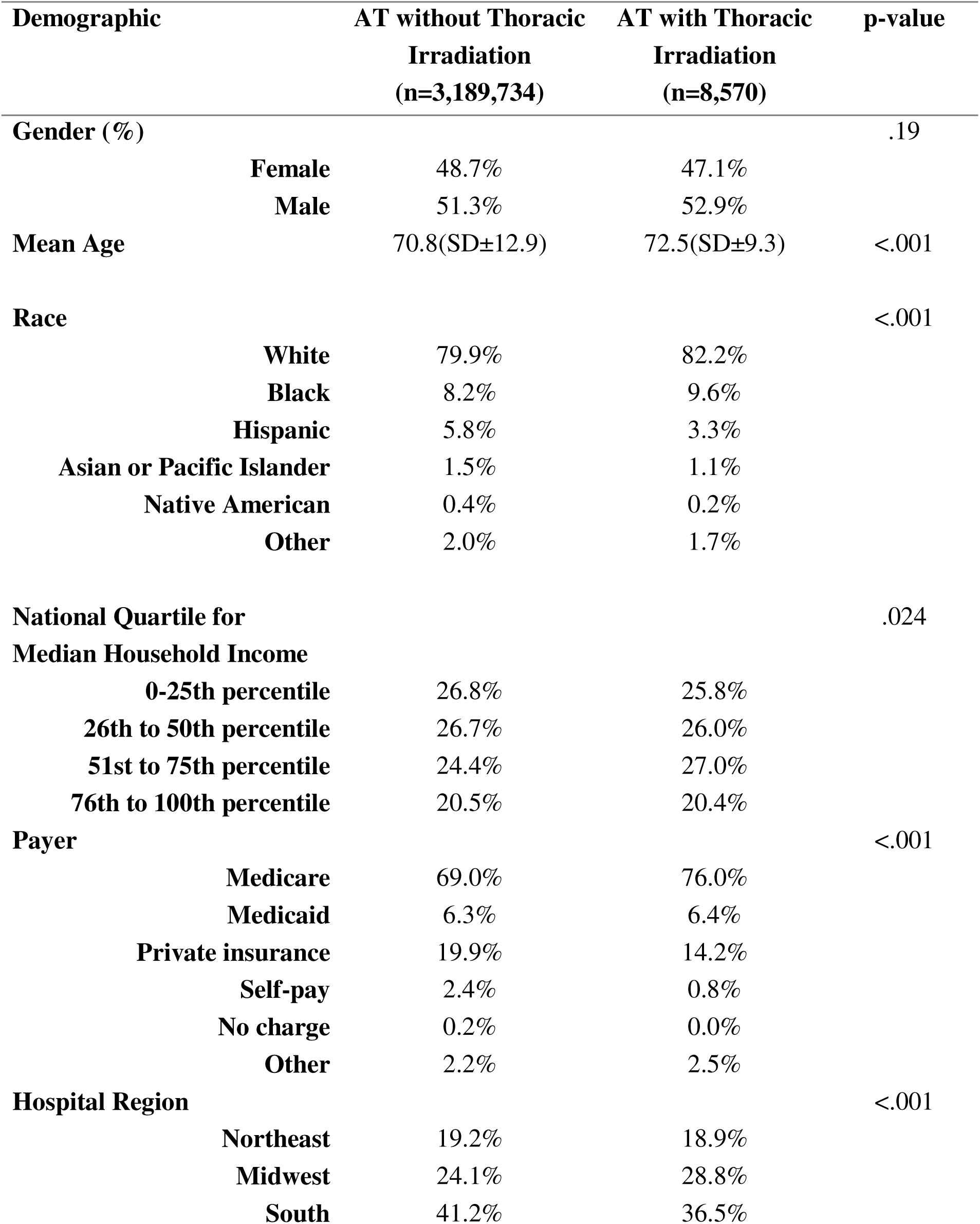
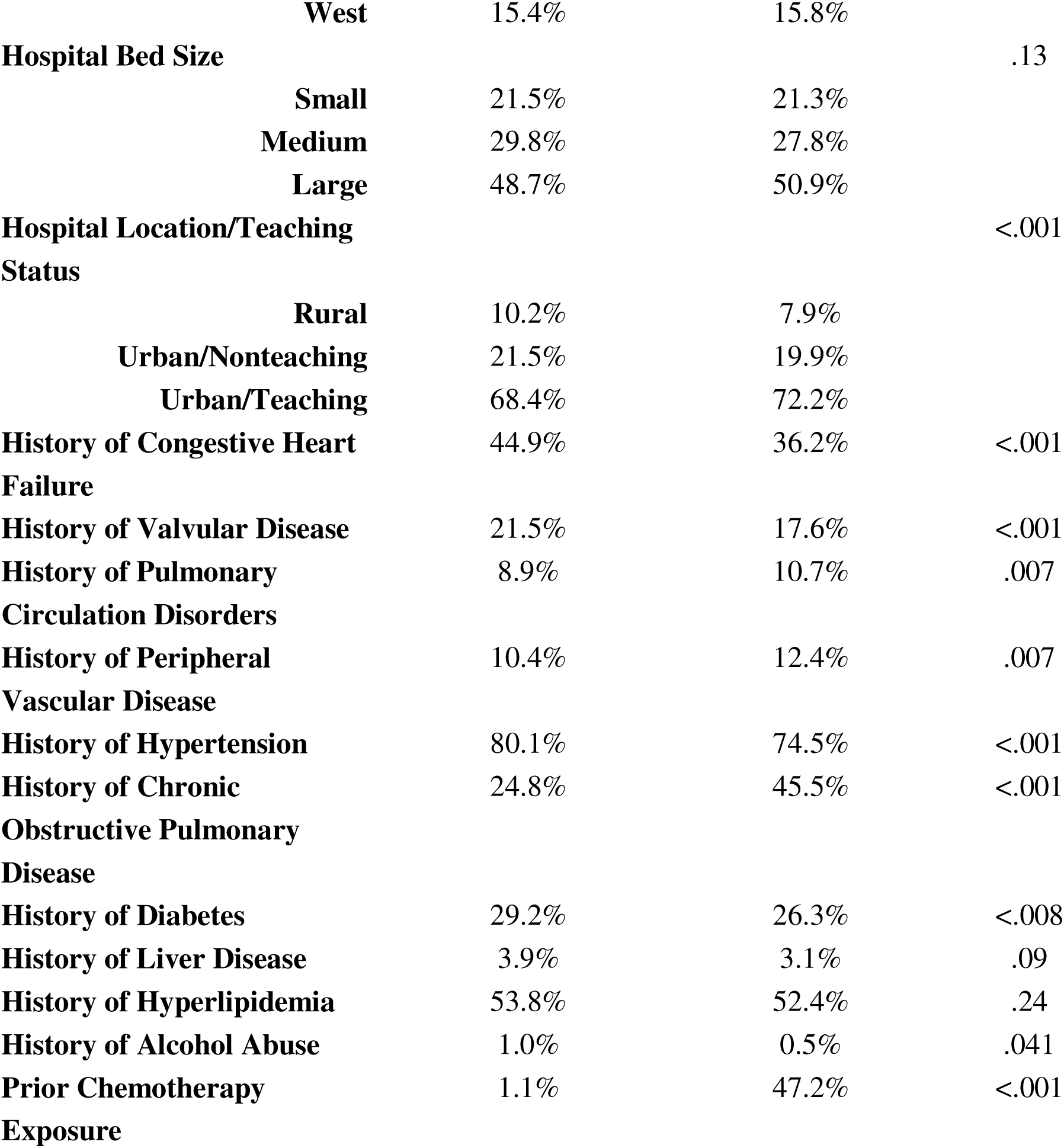
Baseline Demographic and Hospital Characteristics of Patients Admitted for Atrial Tachyarrhythmias (AT) Prior to Propensity Score Matching.

Clinically, patients with prior radiation had lower rates of congestive heart failure (36.2% vs 44.9%, p<0.001), valvular disease (17.6% vs 21.5%, p<0.001), and hypertension (74.5% vs 80.1%, p<0.001), but markedly higher rates of chronic obstructive pulmonary disease (45.5% vs 24.8%, p<0.001) and prior chemotherapy exposure (47.2% vs 1.1%, p<0.001) [Table 1]. Among other comorbidities, diabetes (29.2% vs 26.3%, p=0.008), peripheral vascular disease (10.4% vs 12.4%, p=0.007), and alcohol use disorder (1.0% vs 0.5%, p=0.041) differed significantly between groups, whereas liver disease (3.9% vs 3.1%, p=0.09) and hyperlipidemia (53.8% vs 52.4%, p=0.24) displayed nonsignificant differences [Table 1].

**Table 1**. Baseline variables including age, gender, race/ethnicity, median household income quartile, insurance type, hospital region, hospital size, teaching status, and comorbidities were compared between patients with and without prior thoracic radiation

Among patients with prior thoracic malignancy, lung/airway cancer accounted for 63.7% of cases, followed by breast (23.1%), esophagus (5.9%), and other sites (≤3% each) [Table 2].

**Table 2:**
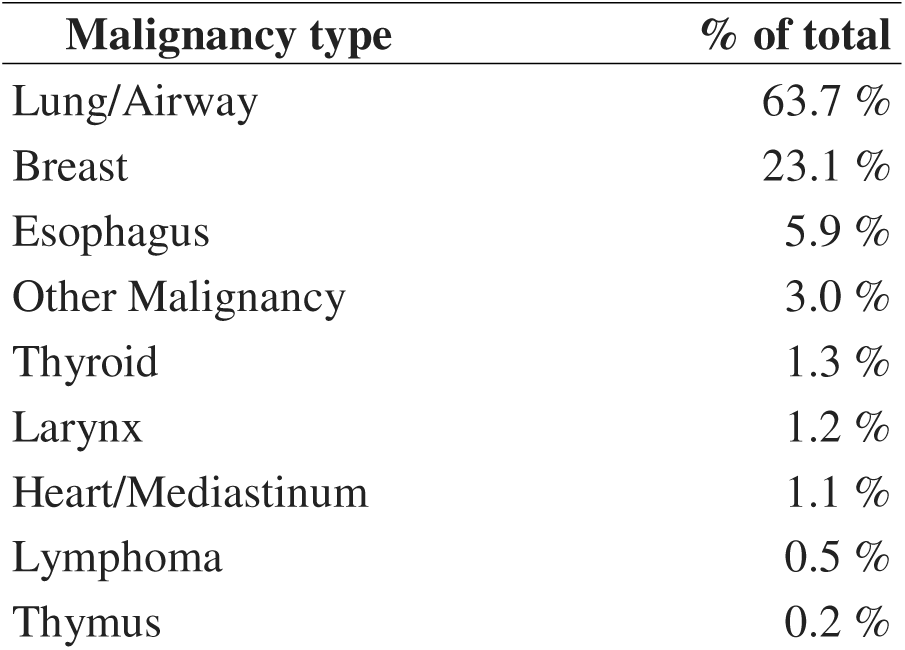
Breakdown of malignancy types.

| **Malignancy type** | **% of total** |
| --- | --- |
| Lung/Airway | 63.7 % |
| Breast | 23.1 % |
| Esophagus | 5.9 % |
| Other Malignancy | 3.0 % |
| Thyroid | 1.3 % |
| Larynx | 1.2 % |
| Heart/Mediastinum | 1.1 % |
| Lymphoma | 0.5 % |
| Thymus | 0.2 % |

**Table 2**. Distribution of thoracic cancer subtypes among patients with a history of thoracic malignancy admitted for atrial tachyarrhythmias, presented as a percentage of total admissions.

### Primary Analysis

In the propensity-matched cohort, prior thoracic irradiation was associated with nearly twice the odds of in-hospital mortality (adjusted odds ratio [aOR] 1.97, 95% CI 1.17–3.32, p=0.010) and a modest increase in length of stay (+0.30 days, 95% CI 0.05–0.55, p=0.019) [Table 3, Figure 1]. Total costs were not significantly differen**t** (mean difference –$2,537, p=0.202) [Table 3, Figure 1]. Irradiated patients had significantly higher odds of palliative care consultation (aOR 2.60, p<0.001) and documented DNR status (aOR 1.97, p<0.001) but lower odds of acute kidney injury (aOR 0.66, p<0.001) [Table 3, Figure 1]. Rates of ischemic stroke, blood transfusion, vasopressor use, mechanical ventilation, cardiac arrest, and cardiogenic shock were similar [Table 3, Figure 1].

**Figure 1.**
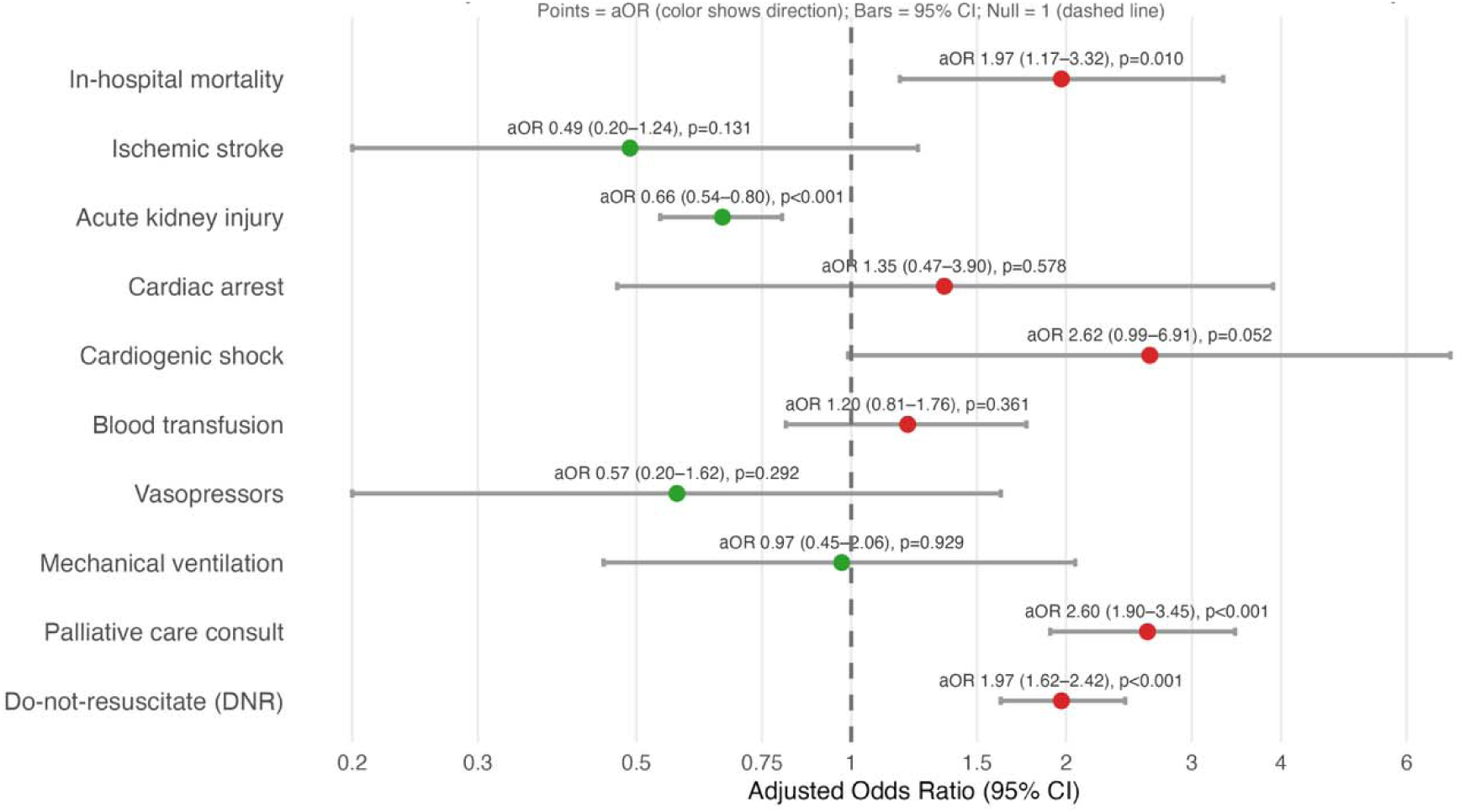
Forest Plot of In-Hospital Outcomes After Atrial Tachyarrhythmia: Prior Thoracic Radiation vs No Radiation (Post-PSM Analysis)

**Table 3.** Multivariable Regression of In-Hospital Outcomes After Propensity Score Matching: Thoracic Irradiation vs No Irradiation in Patients Hospitalized for Atrial Tachyarrhythmias.

| **Outcome** | **aOR/aMR** | **95% Confidence Interval** | **P-Value** |
| --- | --- | --- | --- |
| **Mortality** | 1.97 | 1.17–3.32 | .010 |
| **Total Costs (\$)** | -\$2,536.56 | -\$6,435.00 to<br>+\$1,361.88 | .202 |
| **Length of Stay (days)** | +0.30 days | +0.05 to +0.55 | .019 |
| **Ischemic Stroke** | 0.49 | 0.20–1.24 | .131 |
| **Acute Kidney Injury** | 0.66 | 0.54–0.80 | <.001 |
| **Cardiac Arrest** | 1.35 | 0.47–3.90 | .578 |
| **Cardiogenic Shock** | 2.62 | 0.99–6.91 | .052 |
| **Blood Transfusion** | 1.20 | 0.81–1.76 | .361 |
| **Vasopressors** | 0.57 | 0.20–1.62 | .292 |
| **Mechanical Ventilation** | 0.97 | 0.45–2.06 | .929 |
| **Palliative Consults** | 2.60 | 1.90–3.45 | <.001 |
| **DNR Status** | 1.97 | 1.62–2.42 | <.001 |

aOR - Adjusted Odds Ratio; aMR - Adjusted Mean Reduction; DNR - Do Not Resuscitate;

Interaction testing revealed no effect modification by tumor type for mortality, major complications, length of stay, or total costs (all p-interaction > 0.3). Outcomes with sparse events (e.g., mechanical circulatory support) were non-estimable, but no qualitative interaction trends were observed. Collectively, these findings indicate that the impact of chest radiation on mortality, major complications, and resource utilization was consistent across all intrathoracic malignancy subtypes.

**Table 3**. Clinical outcomes were compared between patients with and without prior thoracic radiation using propensity score–matched, doubly robust multivariable regression. Adjusted odds ratios with 95% confidence intervals and p-values are reported. Interaction terms were incorporated to evaluate potential effect modification by cancer type.

Figure 1. Forest plot displaying adjusted odds ratios, 95% confidence intervals, and p-values from propensity score–matched, doubly robust regression comparing in-hospital outcomes in patients with and without prior thoracic radiation.

### Secondary Analysis

In a sub-cohort limited to patients with either prior thoracic radiation or chemotherapy (but not both), 1:1 propensity score matching produced 7,786 patients (3,893 per group). Radiation-only exposure was not associated with increased mortality or complications compared with chemotherapy-only exposure: in-hospital death (OR 0.83, 95% CI 0.55–1.25), ischemic stroke, acute kidney injury, cardiogenic shock, vasopressor use, mechanical ventilation, palliative consults, and DNR status were all non-significant [Table 4, Figure 2]. The only significant difference was a lower likelihood of blood transfusion in the radiation-only group (OR 0.46, 95% CI 0.34–0.62) [Table 4, Figure 2]. Total hospital costs ( + $562, 95 % CI –$2,453 to $3,576) and length of stay (+ 0.03 days, –0.11 to 0.17) were similar between groups [Table 4, Figure 2].

**Figure 2.**
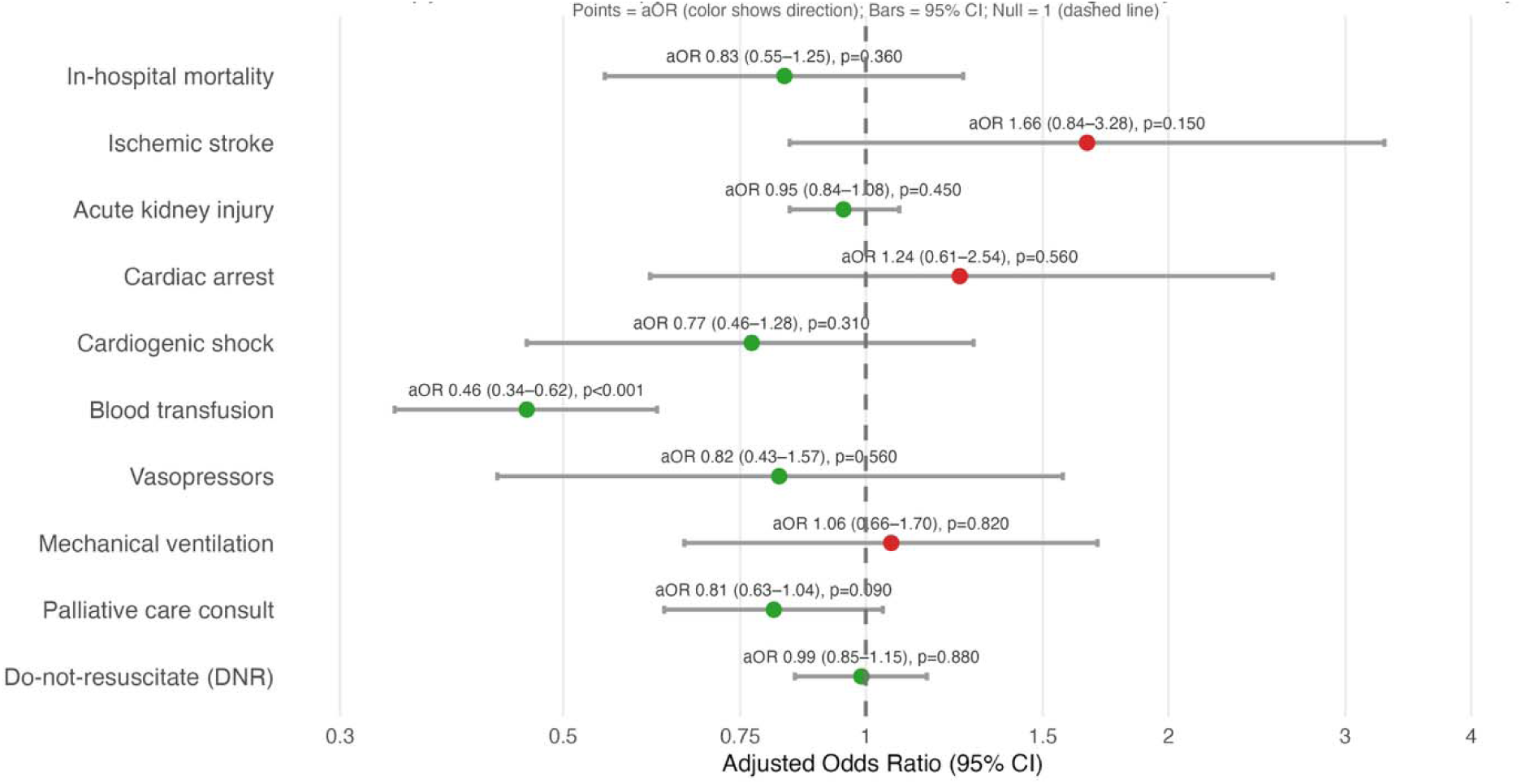
Forest Plot of In-Hospital Outcomes After Atrial Tachyarrhythmia: Radiation Therapy vs Chemotherapy in Patients with Intrathoracic Malignancy (Post-PSM Analysis)

**Table 4.** In-Hospital Outcomes After Propensity Score Matching: Radiation Therapy vs Chemotherapy in Patients with Intrathoracic Malignancy.

| Outcome | aOR/aMR | 95% Confidence Interval | P-Value |
| --- | --- | --- | --- |
| Mortality | 0.83 | 0.55–1.25 | .36 |
| Total Costs (\$) | +\$561.7 | –2452.5 to<br>3575.9 | .72 |
| Length of Stay (days) | +0.03 days | –0.11 to 0.17 | .68 |
| Ischemic Stroke | 1.66 | 0.84–3.28 | .15 |
| Acute Kidney Injury | 0.95 | 0.84–1.08 | .45 |
| Cardiac Arrest | 1.24 | 0.61–2.54 | .56 |
| Cardiogenic Shock | 0.77 | 0.46–1.28 | .31 |
| Blood Transfusion | 0.46 | 0.34–0.62 | <.001 |
| Vasopressors | 0.82 | 0.43–1.57 | .56 |
| Mechanical Ventilation | 1.06 | 0.66–1.70 | .82 |
| Palliative Consults | 0.81 | 0.63–1.04 | .09 |
| DNR Status | 0.99 | 0.85 – 1.15 | .88 |

aOR - Adjusted Odds Ratio; aMR - Adjusted Mean Reduction; DNR - Do Not Resuscitate;

Interaction analyses again demonstrated no effect modification by tumor type for mortality, complications, or length of stay (all p-interaction > 0.13). The only significant interaction was for total hospital costs (p-interaction = 0.036), with heart/mediastinum and breast cancers showing no incremental effect, lymphoma showing a nonsignificant negative trend (p=0.09), and “other thoracic malignancy” tumors showing a significant negative interaction (p=0.002), indicating a relatively lower cost impact compared with non-radiated counterparts.

**Table 4**. Clinical outcomes were evaluated using a doubly robust regression model with propensity score matching, comparing patients with prior thoracic radiation to those treated with chemotherapy. Adjusted odds ratios, 95% confidence intervals, and p-values are displayed

Figure 2. Forest plot displaying adjusted odds ratios, 95% confidence intervals, and p-values from propensity score–matched, doubly robust regression comparing in-hospital outcomes after atrial tachyarrhythmia in patients with intrathoracic malignancy treated with radiation therapy versus chemotherapy.

## Discussion

Overall, this study contributes to the growing body of evidence that both direct and collateral thoracic irradiation contribute to cardiotoxicity, specifically, arrhythmias, and that this particular major cardiac event (MCE) continues to be a leading cause of radiation-induced mortalities in both acute and long-term oncologic care settings [2, 10–12]. In doing so, this study emphasizes that while radiation therapy has become a hallmark for treating breast cancer, small-cell lung cancer (SCLC), non-small cell lung cancer (NSCLC), esophageal cancers, and Hodgkin’s lymphoma, it is crucial to consider dose effects, vulnerable landmarks, co-morbidities, and interaction with other cardiotoxic therapies to optimize oncologic management and mitigate cardiac ramifications.

There has been overwhelming and undisputed risk stratification data regarding both ischemic stroke, coronary artery disease (CAD), and myocardial infarction (MI), as well as atrial arrhythmia incidence. Seminal research from Darby et al. first quantified a 7.4% increase in risk for MCE for every 1 Gy delivered to the heart with prognostic implications lasting 5-20 years post-treatment [1], which was further enhanced by evidence of TRT-related MCE independently predicting all-cause mortality [13]. More recently, findings on thoracic radiation therapy (TRT)-induced atrial arrhythmias show onset may occur as early as 6.7 weeks post-TRT, and the occurrence of cardiotoxicity significantly confer 12 times worse overall survival outcomes (HR 12.1) compared to controls [14], raising concern for increased propensity toward atrial arrhythmogenesis.

While prior research contextualizes our study, this is the first to use a large nationally representative cohort of arrhythmia related hospitalizations to evaluate hospital outcomes among patients with prior TRT using propensity score matching and doubly robust adjustment. Our findings build on Guha et al., who reported increased LOS and hospital costs among cancer patients admitted with AF from 2003–2015 [15]. Using more contemporary data (2016–2022) and including both AF and flutter, we similarly found longer LOS in TRT-exposed patients but no difference in total costs after matching [Table 3, Figure 2]. This suggests that prior radiation is associated with prolonged recovery or monitoring rather than cost-driving interventions, highlighting a distinct utilization pattern in this population.

Most significantly, intensive 1:1 propensity matching and multivariate regression analysis showed that patients with a history of TRT had almost 2 times the odds of in-hospital mortality compared to controls without prior TRT (adjusted odds ratio [aOR] 1.97, 95% CI 1.17–3.32, p=0.010) [Table 3, Figure 1] even when controlling for age, race, insurance type, comorbidities, and hospital factors [Table 1]. Such mortality coincides with prior research from Agarwal et al., who accounted for a significant increase in incidence of atrial fibrillation and (48% vs. 2.4%; p < 0.001) and high-grade AV block requiring pacemaker implantation (20% vs. 9.1%; p = 0.007) in patients with prior-TRT undergoing Transcatheter Aortic Valve Replacement [10]. Combined, the data make a powerful argument that TRT-induced arrhythmia patients are high-risk, growing, and prone to acute decompensation in the hospital setting. Importantly, stratification by tumor type revealed no significant effect modification, further strengthening the generalizability and robustness of our findings.

In radiation-induced cardiac disorders, traditionally, the widely regarded mechanism for such arterial and atrial vulnerability is a sustained anti-oxidant NF-kB targeted cascade, which is triggered by imbalances in reactive oxygen species (ROS) due to radiation [16], resulting in indefinite inflammatory sequelae with upregulation of IL-6-dominant cytokine remodeling of tissues [17, 18]. Dosimetric-volume studies show that these findings hold especially true for pulmonary vein (PV) V5 dose which is significantly associated with atrial fibrillation ([sHR]: 1.04 per mL; 95% CI: 1.01–1.08; P = 0.016) and left circumflex artery (LCX) V35 dose (sHR: 1.10 per mL; 95% CI: 1.01–1.19; P = 0.028) which is significantly associated with atrial flutter [19].

However, this mechanistic framework, while true for ischemic, valvular, and MI radiation-induced events, only partially explains the etiology of radiation-induced arrhythmogenesis, as our findings further explicate. In addition to significant in-hospital mortality mediated by arrhythmias, patients with prior TRT had significantly higher rates of peripheral vascular disease (10.4% vs 12.4%, p=0.007) [Table 1], characterized by fibrotic networks. However, they more abundantly expressed considerably lower rates of congestive heart failure (36.2% vs 44.9%, p<0.001), valvular disease (17.6% vs 21.5%, p<0.001), and hypertension (74.5% vs 80.1%, p<0.001) [Table 1] compared to non-irradiated controls. Such findings are more consistent with anti-fibrotic mechanisms in which an increase in ventricular and atrial conduction velocities is mediated by upregulation of the NaV1.5 ion channel as well as Notch stem cell re-activation [20]. As a result, this further stratifies arrhythmogenic irradiated patients into a complex and vulnerable population, given that typical imaging markers of fibrosis, such as left atrial appendage volume, that are strong predictors of reduced survival, will likely fail to screen highly progressive remodeling of the cardiac electrical circuitry and prevent acute arrhythmogenic decompensation [21].

Under-detection and, thus, increased likelihood of serious arrhythmias among patient with prior TRT is further supported by significantly higher odds of palliative care status (aOR 2.60, 95% CI 1.90–3.45, p<0.001) [Table 3, Figure 1] and documented Do Not Resuscitate (DNR) status (aOR 1.97, 95% CI 1.62– 2.42, p<0.001) [Table 3, Figure 1] in irradiated patients compared to controls. The findings emphasize that once these patients are admitted to an arrhythmia-related event, they reach a clinical threshold in which interventions prove to be futile, necessitating end-of-life care.

While our primary matched analysis demonstrated increased mortality and complications among patients with prior thoracic radiation compared with non-irradiated controls, additional analyses were performed to address chemotherapy as a potential confounder. In a sub-cohort limited to patients with prior thoracic radiation or chemotherapy (but not both), 1:1 propensity score matching yielded 7,786 patients (3,893 per group). Radiation-only exposure was not associated with increased mortality (OR 0.83, 95% CI 0.55– 1.25, p=0.36), ischemic stroke, acute kidney injury, cardiogenic shock, or other major complications compared with chemotherapy-only exposure [Table 4, Figure 2]. Resource utilization was similarly unchanged, with no difference in total hospital costs (+$562, 95% CI –$2,453 to +$3,576, p=0.72) or length of stay (+0.03 days, –0.11 to +0.17, p=0.68) [Table 4, Figure 2]. The only significant finding was a lower odd of blood transfusion among radiation-only patients (OR 0.46, 95% CI 0.34–0.62, p<0.001), consistent with the myelosuppression typically associated with chemotherapy [Table 4, Figure 2]. These findings are supported by pre-existing literature on cardiotoxicity associated with many chemotherapy drugs [17, 18, 22–25], which predispose this sub-population of patients to similar MCEs as irradiated patients. Crucially, by demonstrating no excess risk when radiation is compared directly with chemotherapy, our analysis shows that chemotherapy was a major confounder in unadjusted comparisons and that its exclusion strengthened, rather than weakened, the association between prior thoracic radiation and adverse outcomes. This supports the conclusion that the elevated mortality and complication rates in our primary analysis are not merely artifacts of treatment intensity or cancer-related morbidity but also reflect the independent cardiotoxic impact of thoracic irradiation. Accounting for chemotherapy exposure and using doubly robust propensity score adjustment therefore enhances confidence in the validity of our findings and underscores the importance of considering prior radiation as a distinct risk factor in the inpatient setting.

This study has several limitations. Some findings could not be fully explained, such as the significantly longer length of stay in post-TRT patients compared to controls (+0.30 days, 95% CI 0.05–0.55, p=0.019) [Table 3, Figure 1] despite no difference in hospitalization costs (–$2,537, 95% CI –$6,435 to +$1,362, p=0.202) [Table 3, Figure 1]. Similarly, decreased odds of acute kidney injury (aOR 0.66, 95% CI 0.54– 0.80, p<0.001) [Table 3, Figure 1], diabetes (29.2% vs 26.3%, p=0.008) [Table 1], and alcohol use disorder (1.0% vs 0.5%, p=0.041) [Table 1] remain unexplained and may reflect residual confounding, suggesting that a composite comorbidity index could better capture cumulative burden. Methodologically, reliance on administrative coding introduces potential misclassification bias, and the study lacked cancer-specific variables, radiation dose, and laterality data, which are factors known to influence toxicity, particularly with higher dose to the LAD or left ventricle [26]. Finally, our analysis was limited to inpatient outcomes and did not assess post-discharge complications, late toxicities, or long-term survival.

In light of our study’s findings, patients with prior TRT should be re-stratified as clinically high-risk patients with the potential for severe arrhythmia-mediated complications that require early, increased, and routine surveillance even after radiation treatments. In addition to regular EKGs and advanced Holter monitoring, there is evidence that preliminary rhythm monitoring with heart rate (HR) and heart recovery rate (HRR) provides an accessible measurement that accurately characterizes autonomic dysfunction, a precursor to arrhythmias, and is linked to acute arrhythmia manifestations [27, 28]. Groarke et al. found that prior TRT patients have over five times the odds of higher HRR (aOR 5.32), which was significantly associated with independently predicting an increase in all-cause mortality within 3 years [27].

Furthermore, Gomez et al. suggest that transient changes in biomarkers such as BNP, which remain elevated at first follow-up [29], may provide insight into myocardial stress and warrant early onset of arrhythmogenesis and more intensive monitoring. As such, both propose a non-invasive and timely approach to monitoring severe arrhythmia events.

## Conclusion

Among U.S. hospitalizations for atrial fibrillation or flutter, prior thoracic radiation was linked to higher in-hospital mortality after propensity matching and robust adjustment. Length of stay was slightly longer without increased charges, suggesting extended monitoring rather than procedural intensity. These findings highlight the need for early rhythm surveillance, individualized anticoagulation, and cardio-oncology involvement for this high-risk population.

## Acknowledgment of Financial Support

This study received no external funding.

## Disclosure Statement

The authors declare no conflicts of interest.

## Statements and Declarations

### Funding

The authors did not receive support from any organization for the submitted work.

### Competing Interests

On behalf of all authors, the corresponding author states that there is no conflict of interest.

### Ethics Approval

This study used a publicly available, de-identified database and did not involve any direct patient contact or intervention. Therefore, ethics approval and informed consent were not required.

### Data Availability

The data used in this study were obtained from a publicly accessible online database. Further details are available upon request.

### Author Contributions

Aren Singh Saini and Sahil Ghay contributed equally to this work and are recognized as co–first authors. Both authors had full access to all data and take joint responsibility for the integrity and accuracy of the analysis. Study conception and design were led by Aren Singh Saini and Sahil Ghay. Data acquisition and analysis were performed Aren Singh Saini and Sahil Ghay. The first draft of the manuscript was written by Aren Singh Saini and Sahil Ghay. Baneet Kaur, Shalina Chithriki, Pooja Ghay, Raksha Narasimhan, Kayla Samimi, Isabella Dreyfuss, Roopin Pal Singh, Brandon Mahal, and Crystal Seldon Taswell provided critical supervision, manuscript review, and editorial support. All authors read and approved the final manuscript.

## Notes

### Competing Interest Statement

The authors have declared no competing interest.

